# The Viability, Necessity, and Acceptability of Text Message Group Therapy Among Low-Income Racial-Ethnic Minority Mothers: A Qualitative Study

**DOI:** 10.1101/2024.10.03.24314851

**Authors:** Ilang M. Guiroy, Brianna I. Caicedo, Roya Nouri, Eric Kuhn, Randye J. Semple

**Author notes:** **Correspondence**: Correspondence about this article should be addressed to Ilang M. Guiroy, MD, Child and Adolescent Psychiatry Department, School of Medicine, Stanford University, 401 Quarry Rd, Palo Alto, CA 94305, United States of America.

## Abstract

**Purpose:** This qualitative focus group study conceptually explored the viability, necessity, and acceptability of a proposed text message group therapy (TMGT) intervention as a psychotherapy delivery format among racial-ethnic minority, low-income peripartum mothers.

**Method:** Three semi-structured, virtual focus groups facilitated in-depth interviews with 12 adults who gave birth in the previous 24 months, self-identified as a racial-ethnic minority, had at least mild depression (PHQ-9 >5), participated in a government program requiring low-income (e.g. Special Supplemental Nutrition Program for Women, Infants, and Children [WIC]), and were fluent in English. Recruitment, data collection, and analyses were performed concurrently. This allowed theme saturation (no emergence of new themes) to determine the sample size of 12 participants. The qualitative analysis framework was Grounded Theory. Krippendorff’s alpha was set to > 0.8 for interrater reliability.

**Results:** Ten of the 12 participants were willing to engage in the proposed TMGT. Participants reported a need for psychotherapy tailored to motherhood that was not met by their current support system or healthcare providers, which the proposed TMGT could meet. While all participants communicated via text messaging about motherhood and baby care, they varied in terms of how much. Text messaging has the unique strength of being resilient to interruptions by young children and allowing for multitasking. Text messaging can be a less emotionally stimulating form of communication for an often overstimulated parent. Some mothers found it easier to share their emotions via text message, whereas others found it more difficult. Drawbacks include that information could be lost (increasing frustration or making it possible to hide their true feelings), conversational turns could take too long, and keeping track of the conversation and mobile devices could increase the mental load.

**Conclusion:** These racial-ethnic minority, low-income mothers with peripartum depression expressed a strong need for a safe space to share their lived experiences of motherhood, receive emotional support, and obtain high-quality information about motherhood and baby care tailored to their situation. The proposed text message group therapy is viable, acceptable, and necessary and could meet the needs of many women and increase access to group psychotherapy for this important, underserved population.

---

Many mothers struggle to access psychotherapies for peripartum depression (Goodman, 2009). This is especially true for low-income racial-ethnic minority mothers (Abrams et al., 2009; Bellerose et al., 2022). Digital peer support communities are flourishing in this service gap. For more than two decades, online communities and social media have enabled people to exchange advice, support, and information (Drentea & Moren-Cross, 2005 and Barak 2008). Peripartum mothers have high and sustained engagement with these platforms (Bartholomew et al., 2012). However, it is not well understood how healthcare systems might use social media to expand beyond peer support and how to increase access to psychotherapy led by trained therapists. The engagement of peripartum mothers on social media for emotional support, interpersonal connection, and psychoeducation has prompted this exploration of the clinical potential of digital platforms to increase access to psychotherapy for peripartum depression.

Between 2017 and 2020, the global prevalence of peripartum depression increased from 17% (Shorey et al., 2018) to 31% (Basu et al., 2021). Untreated peripartum depression can be detrimental to mothers’ mental and physical health (Slomian et al., 2019). The effects of depression can extend beyond mothers, with lasting negative consequences for infants, children, and intimate partners (Slomian et al., 2019). Psychotropic medication (Molyneaux et al., 2014) and psychotherapy (Goodman, 2019) are effective treatments for peripartum depression. Often, mothers and providers prefer psychotherapy first, given concerns such as medication side effects on breastfeeding babies or maternal sedation, which could complicate or compromise the 24-hour care needed by babies (Goodman, 2019).

Despite the existence of efficacious treatments for peripartum depression, many mothers struggle to access care. Not only are low income racial-ethnic minority mothers at an increased risk of developing peripartum depression, they also have less access to care (O’Hara & Swain, 1996). Mothering in poverty shaped women’s experiences of depression. These included ambivalence (“I wasn’t prepared for this baby”), caregiving overload (“I need a break”), juggling (“everyone depends on me”), mothering alone (“I really don’t have any help”), and real-life worry (“it’s not safe here”). Barriers to accessing care include stigma against mental illness, fears of being a “bad mother” or “crazy,” long wait times, lack of providers, demands of domestic labor (childcare, managing household logistics, and household tasks), inability to take time off from work, and cost of treatment, and lack of appropriate coverage by insurance. (Abrams & Curran, 2009)

The widespread adoption of technological advances has increased access to online, group conversations. In the past, users were only able to access online platforms using bulky, corded desktop computers. Currently, user-friendly interfaces are easily accessed via mobile devices (e.g., smartphones, tablets, and laptops), which have increased access to online group conversations. According to recent survey, 97% of American adults of childbearing age owned smartphones. Of Americans earning less than $30,000 annually, 79% own a smartphone. Demographically, 84% of Black adults, 91% of Hispanic adults, and 97% of Asian English-speaking adults own smartphones (Pew Research Center, 2024).

Many peripartum mothers build extensive emotional peer support networks via the internet, mobile devices, and social media (Holtz et al., 2015; Johnson, 2014, 2015; Madge & O’Connor, 2006; Stana & Miller, 2019). Despite the increased responsibilities associated with caring for newborns, peripartum mothers report increased social media use to build and maintain a mix of strong- and weak-tie social support networks (Bartholomew et al., 2012). New mothers have a high need for health information and have less time to seek it. Digital support networks on social media for peripartum mothers consist of strong- and weak-tie networks. Strong-tie networks often consist of family members and close friends, are characterized by long personal histories, and who are readily available to provide support. Weak-tie networks consist of people characterized by their availability only in certain circumstances and whose primary role is to share information, differing perspectives, or provide encouragement (Bartholomew et al., 2012). Notably, these support networks can provide entertainment, allow mothers to share information, and promote social interactions during an isolating phase of life (Holtz et al., 2015). In fact, having weak-tie relationships with other mothers online may present an opportunity to allow for more honest revelations of their mothering experiences (Schoenebeck, 2021). Many low-income women seek information and reassurance from online sources such as websites and group forums. Women with high school degrees or less education appear to prefer peer lived experiences as sources of baby care and motherhood information (Guerra-Reyes et al., 2016). This is an important nuance when building trust with this population, since the values of healthcare system emphasize expert opinion or research-backed information.

Text message therapy involves written conversations between a psychotherapist and a patient via a secure platform. This treatment modality is similar to video telepsychiatry, which also directly connects therapists and patients in real time. Mounting evidence suggests that text message therapy can improve clinical outcomes. A large (*N* = 297) multicenter randomized controlled trial (RCT) found that cognitive-behavioral therapy (CBT) for depression delivered by a licensed psychotherapist via text messages decreased depressive symptoms more than those receiving usual care from their primary care provider, with benefits being maintained eight months following the intervention (Kessler et al., 2009). A meta-analysis suggested that text message individual psychotherapy can lead to significant and sustained improvements and was equivalent to, but not superior to, face-to-face or telephone counseling (Hoermann et al., 2017). Despite increasing research on text message therapy, significant knowledge gaps remain regarding text message therapy in group settings.

Text message group therapy (TMGT) is the healthcare system analog of processing and psychoeducation among weak-tie networks on social media. Our definition of TMGT consists of group therapy led by a licensed psychotherapist with three to six patients, usually with a shared mental illness or life challenge. Group therapy sessions consist of live, written message exchanges conducted via a group text message platform that meets patient information privacy standards. Typically there is a regular meeting time and designated session start and end times. The type of therapy orientation is variable, ranging from psychodynamic psychotherapy to CBT.

TMGT is a promising, understudied approach to increasing access to care. It also has the potential to be deployed into established virtual communities on social media platforms. This would require necessary collaboration with social media companies. The platforms would need to be adjusted to comply with the legal and regulatory standards of healthcare systems, such as patient information privacy. Zoom and Microsoft Teams are examples of commercial communication and collaboration platforms that have made this leap, resulting in widespread adoption by healthcare systems to communicate with patients or within treatment teams. Many healthcare providers communicate with each other through group chats. Although not patient-facing, patient privacy secure text message platforms such as Microsoft Teams, TigerConnect, and Voalte have also been widely adopted by many healthcare providers. Popular electronic medical records, such as Epic, have chat functions for healthcare providers. This feature could be expanded to include patients.

Evidence-based psychotherapies would need to be adapted to the TMGT format. One randomized controlled non-inferiority trial (*N* = 179) found that CBT delivered by a licensed psychotherapist via a group chat format to women (97.4%) and men (2.6%) with bulimia nervosa was less effective in producing abstinence from binge eating and purging than CBT delivered face-to-face at the end of treatment. However, this difference mostly disappeared by the 12-month follow-up. The authors concluded that CBT delivered in a TMGT format was an efficacious intervention, although the trajectory of recovery may be slower than in similar interventions delivered in a face-to-face format (Zerwas et al., 2017). A controlled naturalistic study (*N =* 114) of patients discharged from inpatient psychiatric hospitalization found decreased psychological distress and high retention after discharge from inpatient psychiatry hospitalization--only 11 patients were lost to follow up (Golkaramnay et al., 2007). TMGT has been found to be feasible and acceptable among young cancer survivors (Lang et al., 2020) and women with sexual dysfunction (Hucker & McCabe, 2014).

However, it remains unknown whether low-income racial-ethnic minority mothers would find TMGT necessary and acceptable. Low-income racial-ethnic minority mothers are of particular interest, as they face additional barriers to gaining access to treatment. Intersectionality compounds access issues. For the present study, low-income was defined by participating in government assistance programs such as the Special Supplemental Nutrition Program for Women, Infants, and Children (WIC), given that income levels vary nationally.

## Aim of the Study

The present study aimed to assess the viability, necessity, and acceptability of proposed TMGT for low-income racial-ethnic minority peripartum women. Intervention development in the future would then build on this foundation. This was an exploratory study and may be the first of its kind. Therefore, a qualitative approach was chosen to obtain in-depth information about peripartum mothers’ perceptions of a proposed TMGT. The present study sought to (a) explore the viability and necessity of text message group therapy among low-income racial-ethnic minority mothers and (b) establish whether text message group therapy would be acceptable to this population.

## Method

### Participants

The final sample consisted of 12 participants, primarily between the ages of 25-34 years old, and included racial-ethnic minority women. All were high school graduates or higher. The majority were not employed outside of the home, and most (75%) having self-reported incomes less than $31,000. All participants received government assistance and had between one and four children (see Table 1 for demographic characteristics).

### Inclusion and Exclusion Criteria

To be included, participants must (a) have given birth in the last 24 months, (b) be at least 18 years of age, (c) have scored 5 or greater on the PHQ-9 (Kroenke et al., 2010), (d) be fluent in English, (e) have internet access, (f) be participating in any government assistance program for low-income individuals (e.g. Special Supplemental Nutrition Program for Women, Infants, and Children [WIC]), and (g) self-identify as a racial-ethnic minority (American Indian or Alaskan Native, Asian, Black or African American, Hispanic or Latino, Native Hawaiian, or other Pacific Islander, including when mixed with White or European American) on screening to enter the study. Notably, this screening racial-ethnic data to be included in the study was gathered prior to consent and thus was not recorded. As part of the study, participants provided their demographic information via survey. However, half of the participants selected “declined to state” for age and race.

Exclusion criteria were (a) current suicidal ideation, (b) imminent danger to self or others, (c) currently in a manic episode, or (d) currently psychotic. Of 198 potential participants, 12 participated in three virtual focus groups of three to five participants each (See Figure 1). Participation in government assistance programs was chosen as the indicator for low-income rather than household income, as the costs of living varies nationally.

### Focus Group Design and Rationale

This study is exploratory, and to our knowledge, the first of its kind to explore mothers’ views on a proposed TMGT. We chose in-depth, semi-structured focus groups to allow for rich descriptions, contextual information, and an understanding of the common lived experiences of perinatal depression in racial-ethnic mothers facing economic constraints.

Focus groups were conducted online via Zoom video conferencing to eliminate travel costs, allow for geographic diversity, and reach underrepresented populations (Rupert at al., 2017). Each focus group consisted of three to five participants. The focus groups started with an orientation, including a request to keep their cameras on and participants to unmute themselves to allow for spontaneous conversation. The focus group leader (a licensed psychiatrist) provided an overview of the discussion topics. She explained that her role in the group was to guide the structure of the conversation and manage the available time. The focus group leader (IMG) is an experienced group therapy facilitator, skilled in moderating difficult discussions, and maintaining an emotionally and culturally safe environment. She also has clinical expertise in maternal mental health and in serving low-income and primarily ethnic-racial minority communities both in rural and urban settings. Consistent with Grounded Theory, the focus group leader used a short interview guide with open-ended questions. The focus group questions addressed (a) sources of emotional support and health information, (b) modes of communication with support networks, (c) experiences with mental healthcare, and (d) text message group therapy. TMGT was described as group therapy with mothers of babies struggling with stress and low mood, led by a licensed psychotherapist, and with all communication through text messages.

### Procedures

The study procedures were approved by University of Southern California. Potential focus group participants were recruited nationally via social media advertisements posted on social media and locally via community flyers in clinics, laundromats, and grocery store billboards. Participants also included women who had participated in a national web-based survey seeking to understand their opinions as to the viability, necessity, and acceptability of a potential text message group therapy intervention. Eligible participants completed an informed consent form and provided their availability. Participants were recruited between October 2020 and March 2021, with focus groups conducted between November 2020 and March 2021. Focus groups were held via the Zoom video conferencing platform. Professional transcription services transcribed the audio recordings. The focus group leader deidentified the transcripts. Grounded theory thematic analysis and focus groups were conducted concurrently. Recruitment was halted when focus group themes were saturated and no new themes emerged. Participants received $25 Amazon gift cards for their time.

### Analytic Plan

Grounded Theory is an established qualitative approach in healthcare research (Chapman et al., 2015). Grounded Theory (Charmaz, 2012) was selected for qualitative analysis because this method allows for theoretical insights to arise directly from the focus groups. This grounds the research in the voice of the community that it is meant to serve. The research team went through iterative systematic cycles to inductively elaborate, deduce, and verify the data.

Theme saturation determines the sample size (Francis et al., 2010). Theme saturation occurs when no new themes emerge from the data. Consistent with Grounded Theory, recruitment, focus groups, and code book development occurred concurrently. The themes reached saturation (no new themes emerged) with the third focus ground and recruitment stopped. Krippendorff’s alpha was set to >0.8 for interrater reliability.

The three-person analysis team (xx blinded initials xx) went through iterative and systematic cycles to inductively elaborate, deduce, and verify the data. Two independent coders with established experience in qualitative research iteratively analyzed the transcripts. The third person served as the subject matter expert and tiebreaker. The independent coders memoed and consolidated the memos into themes in a codebook. Field notes and coder countertransference were also considered. The three coders met, discussed the themes, and built a consensus codebook. They repeated the process with every focus group transcript until no new themes emerged. Recruitment and data collection were then terminated. The analysis team finalized the codebook. Once data collection was completed, the two coders independently applied the codes to the transcript using Atlas.Ti 21.2.1 for Mac (ATLAS.ti Scientific Software Development GmbH, 2021). The analysis team then met weekly for several months to discuss the codes, refine the codebook, and independently recode until Krippendorff’s alpha was > 0.8.

## Results

### Theme 1: Willingness to try Text Message Group Therapy

When asked, 10 out of 12 participants found the proposed text message group therapy (TMGT) intervention acceptable and said they would be willing to try it. They described many of the themes discussed in detail below regarding how TMGT may fit into their lives and the needs it could meet. The two negative responses focused on privacy and not wanting to talk when feeling low.

> *“Yeah. I would be willing to give it a shot.”*

> *“I would do something like that.”*

> ***“****I really don’t like sharing to strangers about personal detail.”*

> *“And sometimes when I’m down, I just don’t want to talk.”*

> *“I would just be curious about how it would be run, and what app would be used, just more of the logistics. But I think the ideas and the concepts are there. I just wonder how it’s going to play out, honestly.”*

### Theme 2: I’m seeking to improve my caregiving and motherhood experience

Every participant reported seeking information about motherhood and baby care to improve their caregiving and better manage this phase of life. The psychotherapy aspect of TMGT could meet this need. Sources included individuals they knew in person (e.g., family, spouse, friends), online (e.g., motherhood Facebook groups, momfluencer accounts, motherhood WhatsApp groups), and healthcare systems (e.g., pediatricians, therapists, lactation consultants, postpartum support groups).

The participants reported gaps in their support networks concerning knowledge and emotional guidance, indicating a need for the psychoeducation and psychotherapy aspect of TMGT. Participants sought guidance from websites and social media platforms. These digital communities included strong- and weak-ties, including family, familiar friends, unfamiliar strangers, and anonymous users. The resources included websites participants identified through search engines.

> *“If there was something I didn’t know, I did Google it. Like my baby’s bones, when you pick him up sometimes you hear [the joints] cracking… So, I Googled that, like ‘What does it mean?’”*

Many participants sought digital peer support groups on social media platforms. These included Facebook groups (where multiple users with shared interests communicated by posting questions or comments on a website visible to all members) and WhatsApp group chats (where multiple users could communicate rapidly via written messages). In each focus group, participants brought up seeking guidance on social media about breastfeeding.

> *“I’m part of a mom’s group on Facebook as well. I was having quite a difficult time the first couple of months with my son while I was breastfeeding.”*

> *“[The Facebook peer support] groups help, because I didn’t know nothing about breastfeeding and if I have a question, I’ll ask the admins or whatever, the mods and they’ll talk with me in the messages.”*

### Theme 3: Determining the trustworthiness of a resource

Participants described receiving both helpful and unhelpful guidance from their in-person and digital networks. For in-person networks, participants looked at the advisor’s parenting results or whether the guidance had previously helped. The group and psychotherapy aspects of TMGT could meet the need for helpful guidance.

For their digital networks, participants received guidance from unfamiliar strangers or anonymous users. Participants determined whether this guidance was trustworthy by (a) reading through a wide range of lived experiences, (b) deciding whether the guidance or information fit with what they already knew, (c) whether it intuitively made sense, (d) cross-referencing between lived experiences and websites, (e) whether others had previously followed their advice, and (f) most importantly, whether the same guidance came up repeatedly. They also looked to authority figures, such as Facebook group “admins”, individuals who had successfully raised multiple children or healthcare providers (e.g. therapists, pediatricians, and lactation consultants).

Trust is an important aspect of acceptability in therapy, especially among low-income racial-ethnic minority women, for whom authority figures may not always have acted in their best interest, intentionally or unintentionally. The group aspect of TMGT provides trust as peers share lived experiences. The therapy aspect of TMGT provides trust with an empathic, sensitive, knowledgeable, and experienced therapist. The participants were very interested in having a therapist who could keep the conversation emotionally safe and provide reliable health information.

> *“Because if they were the admins of the group, then they basically know what they’re talking about because [otherwise] people aren’t going to follow them.”*

Only one participant reported that she was concerned about privacy and sharing details of personal life with people she did not know.

### Theme 4: I need a safe space to talk about negative things

Participants spoke about the need for a space to be vulnerable and opportunities to process the experience of motherhood that the psychotherapy aspect of TMGT could provide. They felt isolated and lonely. Most participants reported feeling shamed, judged, or burdensome.

> *“I don’t feel like some people understand when you have issues. So it makes you not want to talk to some people…”*

> *“When I’m having just a frustrating day with my two-year-old… It’s hard reaching out to people because I feel like I’m being repetitive like [another participant] said. And it’s just lonely.”*

These participants yearned to be listened to and witnessed.

> “Sometimes it’s just seeing that someone slows down in their world just to listen and talk to you.”

> *“Venting, and I don’t know, just talking about motherhood pretty much.”*

### Theme 5: Mothers must multitask

Participants described managing multiple demands on their time. The texting aspect of TMGT could meet participants’ need to multitask responsibilities that might otherwise interfere with access to care. This theme emerged in the dialogue and on video. One participant actively contributed to the conversation, while she nursed two children, broke up a sibling fight, cooked lunch, admired a child do the Floss Dance, and answered logistical questions from a partner. Participants noted that text messaging allowed for interruptions because the user could catch up and rejoin the conversation.

> *“I’ll be in the middle of a conversation, and the baby will cry, and my son will need something, so on, and so forth. So, I’m all over the place. Or I’ll just randomly get distracted.”*

### Theme 6: Only other mothers truly understand me

The participants emphasized the importance of peer support. The group aspect of TMGT could meet this need. Specifically, those who experienced motherhood could provide support and guidance more effectively than those who had not experience motherhood. Focus group conversations demonstrated this. Participants frequently seconded other participants’ experiences or opinions, expressed empathy, and provided reassurance to each other. An important accompanying theme was, “Sometimes healthcare providers don’t understand motherhood.”

> *“I just enjoy talking to other moms because you can’t … understand what someone is going through unless you have went [sic] through it yourself… I don’t feel like you fully grasp how much responsibility it may be. But if you’re a mom… we can talk to each other and actually understand how one another feels and maybe we have a voice for each other.”*

### Theme 7: Advantages and disadvantages of in-person communication

Text-message communication did not fully replace in-person communication. In-person communication allowed for physical touch and pooling of resources, such as sharing the domestic labor and childcare burdens. Coders coded both the disadvantages of in-person communication and the advantages of audio-visual and text-message communication. These quotes demonstrate some of the limitations of TMGT and some needs that the participants expressed that TMGT could not meet.

> *“Yeah, there’s some texting… I’m more like hands-on. I need them here with me. I need that hug from people that I love and that I know care about me.”*

> *“I talk on face-to-face because my sister currently is up here with me helping me. So when I go in and have my [second] baby, she’ll be here to keep my own one-year-old.”*

### Theme 8: Advantages and disadvantages of audio-visual communication

Many participants expressed a desire for telehealth appointments that could be provided by the digital aspect of TMGT. For multiple participants, in-person appointment increased the amount of domestic labor, including arranging transportation and the logistical challenges of arranging childcare. Childcare often carried interpersonal or financial costs. Many had to bring their children along with them, which required additional time and effort. These tasks included identifying necessary items; ensuring they were available and functional; packing the a bag, snacks, and toys; transitioning the baby and other children; ensuring the baby and other children were fed, changed, and dressed; getting oneself ready and presentable; keeping track of all the necessary items; securing the living space; managing one’s own and baby’s emotions; arranging for transportation; getting the baby and other children in and out of the transport; the sacrifice of time; and the physical, emotional, and mental effort needed to accomplish all these tasks while under pressure.

> *“Because I already have extreme anxiety, so when I have in-person appointments my brain will scramble through a million excuses to not go… [Telehealth appointments are] a lot easier with lack of childcare, trying to get 100 things done, not wanting to drive the 20, 30 minutes there and back for a 45-minute appointment.”*

While most participants were happy with text messaging their support networks, text messages did not replace audio-visual communication.

> *“I see [adult] people so little that I’m like, ‘Talk to me! Please!’”*

Audio-visual communication had some unique advantages over text-message communication. These allowed for a rapid back-and-forth dialogue, but required more attention. This is one need that participants have that TMGT could not meet.

> *“I don’t really like texting myself just because sometimes when you’re in a phone call or when you’re video chatting somebody, you have the time and you have that person’s attention and they have your attention, so it’s a one-on-one and it’s back and forth.”*

One drawback was that young children often disrupted audio-visual communications. When texting, participants reported that their children may come and quickly lose interest, thus minimizing interruptions. This is a need that TMGT could uniquely meet, given that many participants were simultaneously providing childcare.

> *“Yeah, when the [kids] see you on the phone, they don’t leave you alone.”*

> *“Well, let’s pretend I’m talking on the phone because most of it is literally me going, ‘Child! Child! Child!’ The minute my hand [pantomimes picking up phone], my son is right there wanting to know everything… Yeah, I’m actually surprised he hasn’t come back and forth five times already.”*

### Theme 9: Advantages and Disadvantages of Text-Message Communication

All participants communicated via text messages but varied in how much.

> *“I love texting.”*

> *“I hardly ever use text messaging.”*

For some participants, text-message communication made it easier for them to participate in difficult conversations. They could be more thoughtful and have greater control over the content of the communication. Some participants preferred not to talk about their intense emotional experiences through text messages. Notably, there were nuances in the differences in the acceptability of the text message aspect of TMGT for discussion emotional issues.

> *“When it’s things that are hard to say, I’ll just send [my spouse] a text about it.”*

> *“It’s hard sometimes to just say those things out loud, or it’s hard to face their response immediately. It’s just easier to put a little bit of space, and have some time to think about it, and then compose something and send it off.”*

> *“I actually get to collect my thought and compose it… When I’m talking, when I’m in the heat of the moment, or when I’m upset, or sad, or tired, or stressed, or whatever the big emotion is, I’ll say the wrong thing, or I’ll just start crying… I’ll actually be able to take the time to focus, and ‘Oh, I want to say this, but not this.’ And it gives me the time to actually word things the way I want them worded, instead of just word vomiting on everything.”*

> *“When I’m going through stuff, I don’t want to see it in texts. And it’s irritating texting a whole paragraph and getting a whole paragraph back.”*

Other participants noted that information could be lost. Sometimes messages were lost in the flow of the conversation or went unacknowledged. A text message may not communicate mood state or emotions the way a facial expression or tone of voice do. Some participants reported that they could hide their feelings more easily.

> *“Especially if I’m really going through something or really struggling in the moment, I can’t wait for a text.”*

> *“Versus texting when it’s a sentence, a reply to five minutes later. Another sentence and wait 10 minutes. It’s not my deal.”*

> *“I’ll just say, “I’m okay.” It’s not like if I’m talking to you. You can’t feel me or hear me that I’m exhausted.”*

> *“There were so many [people] messaging that [my message] wasn’t picked up.”*

An interesting advantage of text messaging is that the participants described feeling overstimulated. Participants described text messaging as a less stimulating mode of communication.

> *“Because having people talk to me after I have a whole day of screaming doesn’t make me feel better. It just gives me a migraine.”*

Keeping track of text-message communication and maintaining the device added to the mental load for some participants.

> *“Most of the time I don’t text…There’s just too much going on.”*

> *“I think my phone might be dead right now, because I hardly ever charge it because … Oh, wait, my charger is over there.”*

### Theme 10: Mixed experiences with the healthcare system

Participants expressed a need for the therapy aspect of TMGT. One participant shared her positive experience with mental health care.

> *“I got really lucky this time around. I have an amazing therapist and an amazing psychiatrist. I was against meds for my entire life up until last year. And when it comes to my mental health, my support team is like my cheerleaders. “*

Most participants described well-known barriers, such as lack of appointment availability and insufficient insurance coverage for therapy. Many participants described feeling rushed through appointments and their concerns being dismissed. Multiple participants reported that the care felt impersonal.

> *“Sometimes I felt like the person was hearing me, but they wasn’t really hearing me. I think they was just kind of sitting there to get paid.”*

> *“Because sometimes doctors see [you] as a number. They listen to you, and they forget what you said, and they have to read it on the paper, because they have so many people.”*

> *“Finding a therapist that you connect with… it just felt like talking to a wall… [the therapist] didn’t have any kids. She didn’t have any health issues. So, she was just like, “I can’t help you.” I think that’s a hard thing, finding somebody who you can talk to that understands what you’re going through.”*

Some described being afraid of getting mental healthcare for themselves or their children because child protective services might take away their children. Another participant engaged in mental health care to get custody of her son returned to her.

> *“But when I went and sought mental health for him, they threatened, ‘Oh, well, it sounds like you can’t handle taking care of a child. CPS can handle him for you.’ And I was just like, ‘I’m trying to get my shit together, and you’re threatening to take away my child because I’m asking for help?’ And it was horrible, and awful.”*

### Theme 11: My doctor does not understand motherhood

The participants expressed the need for targeted interventions for peripartum depression, that the therapy aspect of TMGT could meet. Participants reported a lack of education among their healthcare providers regarding peripartum depression and treatments. Many participants considered healthcare providers’ lack of understanding of the realities of motherhood as a barrier to care. They also reported that providers sometimes did not tailor their care to the practical experiences of motherhood.

> *“It feels like no one’s listening sometimes… They can’t relate to what I go through on a daily basis.”*

> *“The doctors tell you that you’ll get over [peripartum depression] eventually… Most doctors don’t understand.”*

> *“Go to the doctor, and say whatever, and he’ll probably give me a sleeping pill…and something for stress. That’s not going to help me with the babies or anything.”*

Healthcare providers who limited understanding of a patient’s situation may have the potential to put patients at risk. One participant described being in an abusive relationship and that her medication side effects had interfered with her ability to respond during a “domestic issue.”

> *“His dad … would come in here and fight with me and throw shoes at me and I wouldn’t have no response. I would just stand there like a zombie. And then my doctor would be like, ‘That’s good. That’s good.’ But people are walking all over me. I’m like being a pushover, you know?”*

### Theme 12: I want more options than medications

Many participants expressed wanting therapy for peripartum depression, a need TMGT could meet. Most of the time, healthcare providers only offered medications. The participants listed concerns about side effects, breastfeeding, and the stigma associated with psychiatric medications.

> *“My general physician, she pressed and pressed and pressed for me to be on medicine. It doesn’t work for me. I’m super sensitive. It makes me sick. It numbs me. I don’t like that.”*

> *“I don’t want to reach out to a doctor because… I don’t want to be extreme with medications and all that. I’m breastfeeding first off.”*

> *“I want to get to the root of it. I want to get down to what’s going on. I want to talk to somebody. And for [the doctor], it just got to the point where, ‘Okay, a bit more. Okay, well, that’s not working.’”*

### Theme 13: Issues related to COVID-19

This study was conducted after most initial shelter-in-place orders were lifted in the United States, but before most of the population had access to vaccines. At that time, the coronavirus disease (COVID-19) death toll was still high and the effect of COVID-19 on pregnancy, breastfeeding, and young children were unclear. Multiple participants reported that the healthcare system focused on treating patients with COVID-19 and their needs appeared to be dismissed.

> *“I feel like every time I’ve been to the doctor, [COVID] overshadowed my needs.”*

> *“COVID is the center of attention. Makes it feel like whatever was going on with you it’s not important, whatever I’m feeling it’s not priority. If I’m feeling down, there’s people that are having it worse because of COVID.”*

Others described additional stressors, including the lack of childcare and being out of work because of COVID-19.

> *“Right now we don’t have a support system because of the whole COVID thing going on. We don’t have childcare at all.”*

> *“Being out of work…, homelessness and all that because of COVID.”*

## Discussion

The present study consisted of focus groups of low-income racial-ethnic minority mothers with at least mild depression. The focus groups explored the viability, necessity, and acceptability of a proposed text message group therapy (TMGT). The proposed TMGT was described as group therapy with other mothers of babies struggling with stress and low mood, led by a licensed psychotherapist communicating via text messages only. Participants reported that the proposed TMGT could meet their need for psychotherapy delivery in a way that was tailored to the realities of motherhood. The majority indicated that they would find the proposed TMGT acceptable.

Regarding the acceptability of the proposed TMGT, participants were overwhelmingly willing to try group therapy by text message (10 out of 12 participants). Participants demonstrated acceptance of the digital relationship aspect of TMGT by describing that they were comfortable giving and receiving information about motherhood and baby care in online spaces, including with unfamiliar or unknown individuals. The group aspect of TMGT provides trust as peers share lived experiences. The therapy aspect of TMGT provides trust with an empathic, sensitive, knowledgeable, and experienced therapist. All participants communicated via text messages about baby care and motherhood, although they varied in frequency. Demonstrating further acceptability, some participants engaged in motherhood special-interest text message groups with other mothers from their virtual weak-tie support networks. These findings are consistent with previous literature, which suggests that mothers gravitate towards groups of weak-tie peers to access diverse motherhood experiences from mothers outside of their social circles (Holtz et al., 2015). Drentea and Moren-Cross (2005) found that mothers sought support from other mothers who shared similar stressors.

TMGT for peripartum depression can address a unique gap. Previous literature on text message therapy for peripartum mothers has primarily studied the perspectives of mothers receiving prewritten automated SMS messages (Broom et al., 2015; Zhou et al., 2022). This study expands on these findings by capturing preferences related to live text message exchanges from low-income racial-ethnic minority peripartum mothers. There were some nuances in the acceptability of TMGT related to discussions of emotionally difficult topics via text message. Some participants found it easier to communicate difficult emotions through text messages. Others preferred in-person or audio-visual communication for emotional conversations. Participants noted that they needed to multitask and provide childcare while receiving healthcare. Interruptions are often a part of caring for babies and small children. A particular strength of the text message aspect of TMGT is its resilience to interruptions. Participants liked being able to drop out and rejoin a conversation as necessary. Participants noted that young children, who tended to frequently interrupt audio-visual conversations, were less likely to interrupt text-message conversations. They described text messaging as a less stimulating form of communication, feeling more tolerable to an often cognitively-physically-emotionally-overstimulated, multitasking mother. TMGT is a form of telehealth that potentially can decrease the labor required to manage household and childcare logistics, make shouldering childcare burden easier, and decrease the time and financial burden of transportation. Text-message communications, however, were not perceived as a replacement for in-person or audio-visual conversations. There were certain needs that TMGT could not meet, such as physical touch, pooled resources (e.g., childcare), and hearing a comforting voice.

Text-message communication had some unique drawbacks. Some participants reported a loss of information. Sometimes, text messages were lost in the flow of conversation. Without the cues of non-verbal communication, participants noted that it was easier to hide their emotions or avoid engaging in certain topics. Waiting a long time to receive a response to their text message was frustrating for some participants. Keeping track of conversations and mobile devices increased the mental load and logistical burden of managing a household. For group text messages, concerns about privacy when talking with unvetted strangers were noted once.

Regarding the necessity of TMGT, participants described gaps in their current support networks and healthcare systems. Participants indicated feeling lonely, which could be addressed by the group aspect of TMGT. Participants described wanting a safe space to be vulnerable and to process the experience of motherhood. Despite potential progress, participants continued to experience challenges in accessing care. They expressed a desire for psychotherapy tailored to the realities of parenting babies and young children while having limited financial, time, and support resources. They sought high-quality, vetted information from other mothers about managing the stresses of motherhood and caring for babies and young children. and looked to their healthcare providers for help. The participants reported access to peer support. However, these participants lacked sufficient access to therapy tailored to accommodate the demands of motherhood--a need that the therapy aspect of TMGT could meet. Only one participant was able to access mental healthcare without difficulty. Among participants who were able to access care, many reported that they felt their care was compromised by the providers lack of understanding of the unique needs of mothers caring for their babies. Multiple participants sought psychotherapy for peripartum depression but were often only offered medication. The proposed TMGT intervention could increase access to peripartum mental healthcare by combining peer support with guidance from licensed therapists, cost effectively, and with fewer logistical restrictions than in-person interventions.

This qualitative study had several strengths. Semi-structured focus groups and Grounded Theory allowed for the collection of in-depth, rich data while preserving the context to uncover common lived experiences. Rather than assuming or telling what mothers wanted or needed, participants were invited to participate in a dialogue with discussions centered on their views. This study brings their voices into the scientific literature. The focus group facilitator shared similar demographics as the participants, which may have enhanced the richness of the information the participants were willing to provide.

This study had limitations common to all focus group designs, Grounded Theory, and qualitative research. Semi-structured focus groups allowed the conversations to be partially driven by the participants, resulting in natural variation in what was discussed in each group. This was addressed by ensuring that the focus group leader asked specific questions in each group. While participants can encourage each other to participate, focus groups may also make it less comfortable to express dissenting opinions. The focus group leader attempted to mitigate this by implementing process group therapy techniques that encouraged the expression of diverse perspectives and opinions. Although Grounded Theory strives to focus on the responses of the participants, data are processed by human coders who may bring their own unconscious biases or pre-existing beliefs into the analysis. To reduce the risk of this bias, multiple coders conducted analyses using an iterative approach and cross checks.

Although the participants were recruited using a variety of methods, they primarily came into the study through social media advertisements. This may have biased the sample towards those who were more comfortable with technology-enabled communication. During the screening for eligibility, all participants self-identified as non-white. However, given that this information was gathered prior to obtaining consent, the participants were asked to provide this information again by a survey prior to starting the focus groups. This time, many declined to disclose this information resulting in incomplete data on the racial-ethnic data. This study focused on a narrow definition of “motherhood,” defined solely as having given birth. This excludes other ways of becoming a mother, such as adoption and surrogacy. This selection criterion was chosen because of the biological component of peripartum depression.

This study was conducted during the acute phase of the coronavirus disease (COVID-19) pandemic. None of the participants expressed concerns about infection, but rather expressed frustration that the healthcare system was overwhelmed and less responsive to their non-COVID-related health needs. Additionally, participants reported losing their jobs, becoming homeless or having less access to in-person support networks.

TMGT appears to be a viable, necessary, and acceptable addition to currently available treatments for low-income racial-ethnic minority mothers. While these findings are important in advancing TMGT as a therapy delivery modality, much more needs to be navigated before real world implementation can be done. A non-exhaustive list of the next steps include: (a) using or modifying an established platform for group chat between patients and therapists, or co-creating a group text message platform by using an iterative, user-centered design that meets patient privacy standards, that is usable, acceptable, and effective, (b) developing protocols to implement rapid responses to imminent danger to self or others, (c) adapting therapy modalities with efficacy to the group text message format, (d) identifying and training therapists, (e) developing protocols for identifying appropriate patients to refer to TMGT (f) implementing TMGT into clinical workflow integration, (g) ascertaining acceptance of TMGT by healthcare system cybersecurity standards, (h) identifying and training IT support staff, and more. It would also be useful to explore the preferences of patients’ and therapists’ regarding the features that need to be included in a TMGT intervention. An iterative, user-centered development of this intervention would increase the likelihood of successful implementation, acceptability, and effectiveness of TMGT. Future research should include data collection from additional cultures and ethnic groups. By collecting diverse perspectives, we can better meet the unique needs of “invisible groups.” This includes perspectives from Native American and Asian mothers. In addition, expanding the traditional definition of motherhood to include surrogacy, adoption, trans parents, and non-birthing parental figures such as fathers, elder siblings, grandparents, and professional caregivers would allow these voices to be heard.

Although efficacious treatments exist, peripartum mothers experience multiple barriers to accessing mental healthcare. This study found that low-income racial-ethnic minority mothers believed that the proposed TMGT was both necessary and acceptable. Peripartum depression impacts millions of families. TMGT could increase access to treatment for a population with limited mental healthcare access and resources, and that are at high risk for peripartum depression.

## Statements and Declarations

### Data Availability

The participants in this study did not provide written consent for their data to be publicly shared. Due to the sensitive nature of this research, supporting data will not be made available.

### Ethics Approval

Study procedures were reviewed and granted exempt status because it was deemed minimal risk by the University of Southern California Institutional Review Board (APP-19-07355, 9/25/2020) and certified that this study was performed in accordance with the ethical standards of the 1964 Declaration of Helsinki.

### Consent

Informed consent for participation and publication was obtained from all participants in this study.

### Author Contributions

Ilang M. Guiroy, M.D. and Randye J. Semple, Ph.D. contributed to the conception and design of the study. Material preparation, data collection, and analysis were performed by Ilang M. Guiroy, M.D., Brianna U Caicedo, B.A., and Roya Nouri, Ph.D. Ilang Guiroy, M.D., Eric Kuhn, Ph.D. and Randye Semple, Ph.D. contributed to the interpretation of the data. The first draft of the manuscript was written by Ilang Guiroy, M.D. and all the authors provided feedback on the manuscript. All the authors approved the final manuscript

